# VITRUVIUS: A conversational agent for real-time evidence based medical question answering

**DOI:** 10.1101/2024.10.03.24314861

**Authors:** Maria Camila Villa, Isabella Llano, Natalia Castano-Villegas, Julian Martinez, Maria Fernanda Guevara, Jose Zea, Laura Velásquez

## Abstract

**Background:** The application of Large Language Models (LLMs) to create conversational agents (CAs) that can aid health professionals in their daily practice is increasingly popular, mainly due to their ability to understand and communicate in natural language. Conversational agents can manage enormous amounts of information, comprehend and reason with clinical questions, extract information from reliable sources and produce accurate answers to queries. This presents an opportunity for better access to updated and trustworthy clinical information in response to medical queries.

**Objective:** We present the design and initial evaluation of Vitruvius, an agent specialized in answering queries in healthcare knowledge and evidence-based medical research.

**Methodology:** The model is based on a system containing 5 LLMs; each is instructed with precise tasks that allow the algorithms to automatically determine the best search strategy to provide an evidence-based answer. We assessed our system’s comprehension, reasoning, and retrieval capabilities using the public clinical question-answer dataset MedQA-USMLE. The model was improved accordingly, and three versions were manufactured.

**Results:** We present the performance assessment for the three versions of Vitruvius, using a subset of 288 QA (Accuracy V1 86%, V2 90%, V3 93%) and the complete dataset of 1273 QA (Accuracy V2 85%, V3 90.3%). We also evaluate intra-inter-class variability and agreement. The final version of Vitruvius (V3) obtained a Cohen’s kappa of 87% and a state-of-the-art (SoTA) performance of 90.26%, surpassing current SoTAs for other LLMs using the same database.

**Conclusions:** Vitruvius demonstrates excellent performance in medical QA compared to standard database responses and other popular LLMs. Future investigations will focus on testing the model in a real-world clinical environment. While it enhances productivity and aids healthcare professionals, it should not be utilized by individuals unqualified to reason with medical data to ensure that critical decision-making remains in the hands of trained professionals.

## 1 Introduction

AI solutions like large language models (LLMs) and LLM-powered Conversational Agents (CAs) can assist health-care personnel in their daily practices, automating repetitive processes and reducing the pace at which they must constantly update themselves to keep up with the flow of information and knowledge [1]. This allows them to focus on the actual interaction with the patient and improve care. Artificial intelligence (AI) agents can use user input, make decisions, and take action to produce answers [2]. Thus, conversational agents (CAs) can mimic human conversations using text or voice, such as Apple’s Siri, Microsoft’s Cortana, or Amazon’s Alexa. LLMs are trained on hundreds of billions of words from diverse Internet sources [3]. Due to this training, they can transform natural language into structured data to perform various language tasks such as translation, summarization, retrieving specialized and relevant information in response to a query, and sentiment analysis, among others [4]. For the last decade, their capacities have expanded beyond basic language processing thanks to the implementation of prompting strategies (specific ways to give instructions) and external tools [4]. LLM-powered CAs like ChatGPT, Gemini or GoogleBard have risen in popularity for personal and professional use, given their ability to carry conversations, answer questions, design follow-up plans, and access information from various topics.

LLMs can be present in multiple medical domains, including consultations, medical charts, diagnosis, treatment plans, and monitoring. For example, a complete understanding of a patient’s history and/or condition can help with clinical management, especially with patients treated by several specialities, resulting in informed actions and decisions. It can also provide treating physicians with best evidence practices retrieved from trusted medical libraries such as PubMed and official clinical guidelines with a click, improving efficiency and precision in any consultation

Both LLMs and CAs have been tested on several medical question-answering (QA) datasets built from medical licensing exams and natural interactions between doctor and patient taken from telemedicine consultations. Popular examples of these QA databases are MedQA [5], MedMCQA [6] and PubMedQA [7]. LLMs performance has surpassed human passing scores in all three datasets [8–10], and some have even surpassed expert performance, such as GPT-4 when using MedQA and PubMedQA [5]. CAs like ChatGPT have also been tested, attaining passing-level performance in the United States Medical Licensing Examinations (USMLE) [3]. State-of-the-art (SoTA) refers to the highest-performing LLMs using a specific QA dataset. For example, the performance for Med-Gemini using MedQA is 91.1% (accuracy) [6]; using PubMedQA, the GPT-4 obtained 81.6% accuracy, in combination with a particular prompting strategy named Medpromptit [7]; the model Med-PaLM 2 had a 72.3% accuracy using MedMCQA [11].

Newer research has focused on fine-tuning the models to create specialized LLMs and CAs for medical applications such as Med-PaLM 2 [11], BioMedGPT [12], InstructGPT [13], Med-Gemini [14], among others. They have also been developed for particular specialities like radiology [15], oncology [16], ophthalmology [17] and dementias [18], to name a few.

We aimed to develop an LLM-powered CA equipped to produce responses to medical queries in different clinical and scientific research fields. In this paper, we present Vitruvius, our first conversational agent and research assistant, along with the steps followed for its creation. We also assess the model’s accuracy compared to a standard QA database.

## 2 Methodology

### 2.1 Development of the model

Vitruvius is a CA powered by multiple LLMs working together to answer clinical questions using information from the internet, research articles, clinical guidelines and medical libraries. Vitruvius has a real-time Retrieval-Augmented Generation (RAG) system. This system consists of two modules: one that retrieves information and the other that generates text based on this information. Retrieval extracts relevant chunks of information. For our system, the retrieval module uses two Application Programming Interfaces (APIs), which refer to computational strategies that allow the software to communicate with an application as humans would [19]), from Google and PubMed, to search and retrieve relevant information from amongst over 37 million biomedical literature references, primary medical guidelines and research. The generation module consists of the LLMs that use the retrieved information as context to generate a text answer to a query.

The baseline LLMs used are in the Generative Pre-trained Transformer (GPT) family due to their SoTA performance in general language reasoning tasks, such as GPT-4o’s 88.7% accuracy using the MMLU databases [20], 90.5% with MGSM [21], and 90.2% when assessed with a HumanEval [22] (available at: https://openai.com/index/hello-gpt-4o/). The system components are explained in the following sections.

### 2.2 RAG and API systems

#### 2.2.1 APIs

Vitruvius uses the Google and PubMed APIs as sources of information. The Google API can generate search engines capable of building restricted searches to find information on specific pages. We used a functionality called SafeSearch to help avoid retrieving information from unsafe web pages. The PubMed API allows for consulting all free access articles available in PubMed. Data in PubMed can be sorted and filtered according to relevance, publication date, authors, and journals. For Vitruvius, searches are filtered by relevance. Each search using either API chooses the 10 most relevant articles to construct an answer to each specific query.

#### 2.2.2 Workflows

To maximize the accuracy of the answers, we created 4 different workflows in which the question in hand is routed (see workflows below). These workflows correspond to the most frequent clinical question types in our experience with focus groups in health care personnel. The routing is done by LLM 1 (Figure 3.), which we named the orchestrator. It classifies each query based on its purpose and source type needed to answer and activates the corresponding workflow. LMM 1 also receives the answers from the other LLMs and builds a final output for the user, acting as a “ judge” of syntactic and semantic quality. The workflows are graphically represented in Figure 1. and described below:

**Figure 1:**
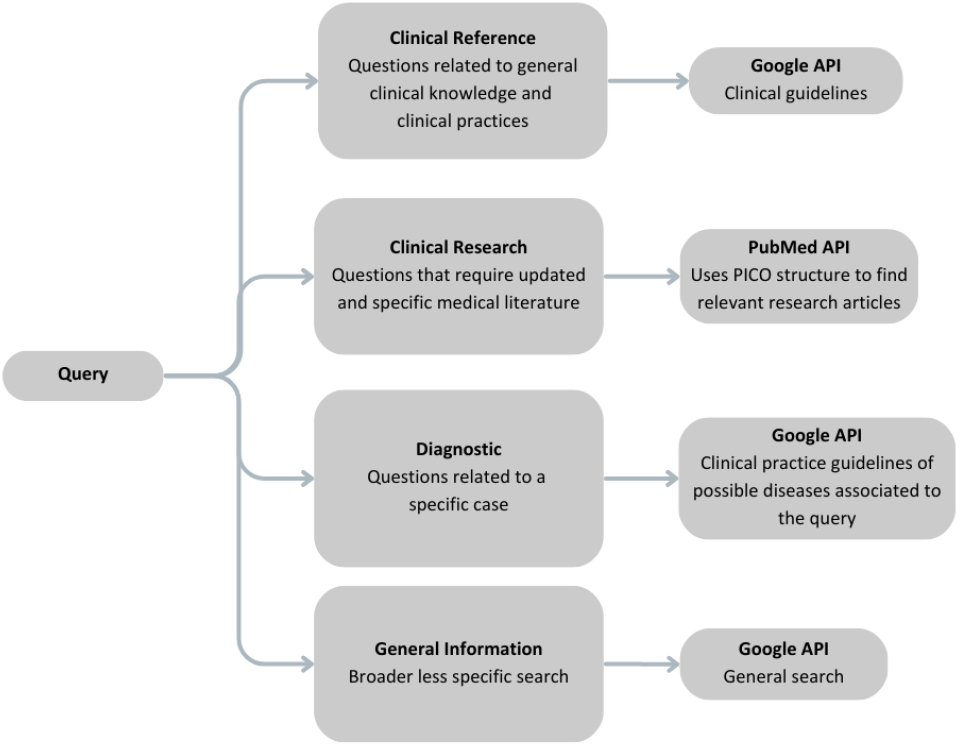
Overview of the different workflows used in the Vitruvius system.

**Figure 2:**
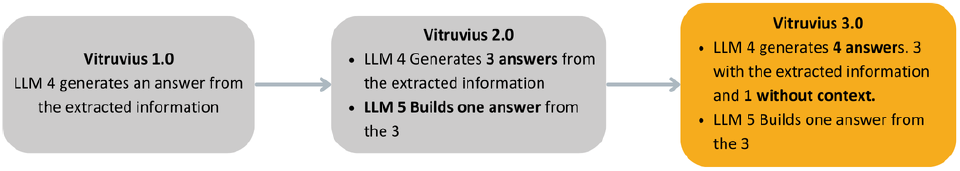
Differences between the three versions of Vitruvius.

**Figure 3:**
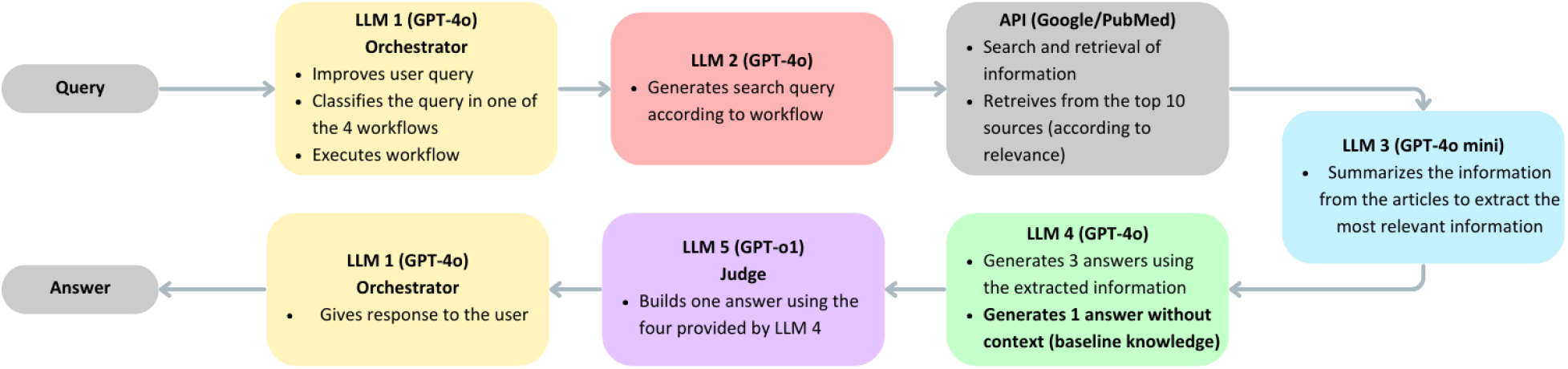
Pipeline used in the Vitruvius system.

1. **Clinical reference questions**: These questions need general medical knowledge, and their responses are provided based on official clinical practice guidelines. This workflow uses the Google API to perform a Google search for the clinical guidelines to answer the question.
2. **Clinical research questions**: Require retrieving research articles to address medical and research queries. This workflow uses the PubMed API. The input question is automatically transformed into a PICO format (Patient, Intervention, Comparison, Outcome) for optimal research results, looking for the appropriate references to answer the question.
3. **Diagnostic questions**: These cases require assistance with diagnostic information. Given a specific case, this workflow uses the Google API to search for clinical practice guidelines to find information about the disease diagnostic protocol associated with the query and produce a response.
4. **General information questions**: These require a broader, less specific search. Thus, this workflow uses the Google API to extract and summarize the key findings.

The system works in English, Spanish, and Portuguese and is instructed to answer in the user’s chosen language. Internal instructions, information search, and retrieval are given in English, and the final response is translated. The model is also instructed to provide literary references, including the title, URL, and APA format, when possible.

Once the orchestrator classifies a query, LLM 2 (Figure 3.) generates the ideal search strategy, which is passed onto the APIs that perform the search. For better and more reliable use, if Vitruvius cannot find information on the query, it will announce it and respond that it has not found information to provide an answer.

#### 2.2.3 Answer generation

Answer generation is divided into three steps. LLM 3, summarizes the information retrieved by the APIs, extracting the most relevant information. Then, LLM 4 generates four answers to the initial query, three using the retrieved information as context and one without context using the LLM’s baseline knowledge. Finally, LLM 5 produces one answer out of the four created by LLM 4. This information is then passed on to the orchestrator, which creates the final response to the user. The complete LLM pipeline is represented graphically in Figure 3.

The process of answer generation, per se, was improved using three experimental iterations before obtaining the most accurate configuration, represented in LLMs 2, 3 and 4. The first version of Vitruvius did not include LLM 5, and LLM 4 generated a single answer from the summarized information. The second and third versions of Vitruvius consider the relevance of the random effect in language generation models, which is expected and produces different responses to the same query. In response, we used this effect in the second version of the model; LLM 4 generated three answers from the retrieved information, and LLM 5 compiled these into one. Finally, in the third and final version, LLM 4 generated four answers, three using retrieved information and one using LLM background knowledge. LLM 5 maintained the same functionality. The differences between each version of Vitruvius is shown in Figure 2.

### 2.3 Dynamic user interface

Vitruvius will be through a custom URL created by the Arkangel AI software platform, which is designed to build custom predictive and generative algorithms tailored for healthcare applications. This platform is the backbone of Vitruvius and enables the LLM integration and core functionalities that make it useful for users in natural clinical settings. Users can input queries on the Arkangel AI platform through a conversational interface that allows follow-up questions and conversation-style interaction between the user and the system (Figure 4.).

**Figure 4:**
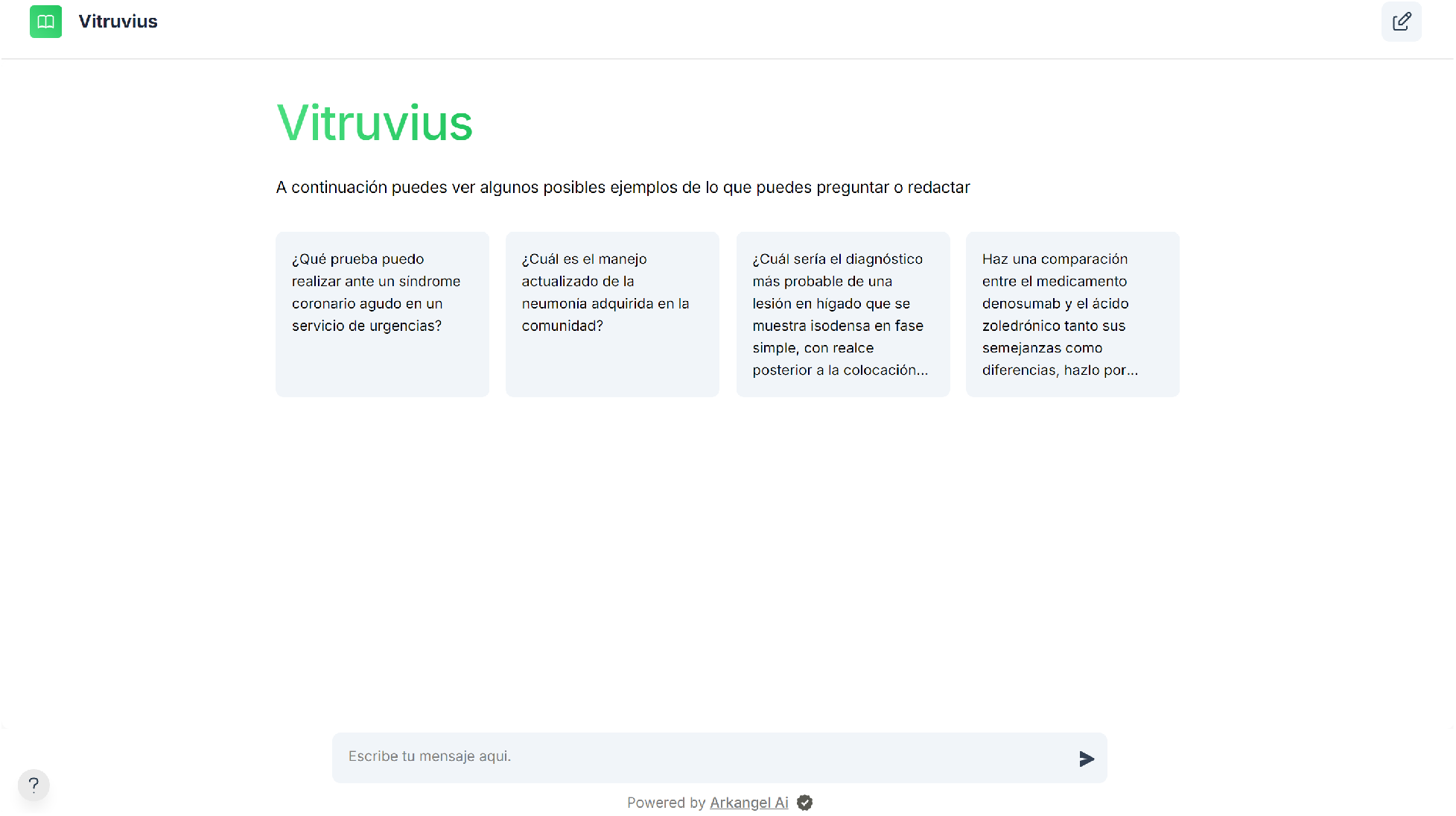
Vitruvius user interface in Spanish (default language)

### 2.4 Model Evaluation

#### 2.4.1 Datasets and Metrics

QA medical datasets are the current benchmark for clinical knowledge and reasoning for LLMs and CAs. For this initial evaluation, we used MedQA. This dataset was first presented in September of 2020. It comprises 12723 multiple choice questions and answers (QA) from the medical licensing exams from the United States (USMLE), Mainland China (MCMLE - 34251 QA) and Taiwan (TWMLE - 14123) in English, simplified Chinese and traditional Chinese [23]. Each set of questions is divided into development, training and testing splits. This dataset includes two main categories of questions: the first, where a single piece of knowledge is required, often requiring one reasoning step, and the second, where a question is asked based on a clinical case; these questions usually require multiple reasoning steps and are more challenging.

For this study, we used questions in English from the USMLE set as it is the most used in evaluations. We only use the test set since Vitruvius doesn’t require training or fine-tuning. The SoTA scores with other LLMs are available at https://paperswithcode.com/sota. The test set from MedQA includes 1273 questions.

We evaluated internal and external variability to thoroughly assess the dataset and model bias and the model’s ability to answer correctly. For internal variability, we will calculate sensitivity/recall 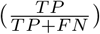 precision 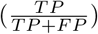 and F1 score 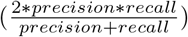 per class (A, B, C, or D). Agreement with standard answers from MedQA will be evaluated with the metric accuracy 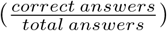 as it is the benchmark metric for SoTA and concordence with Cohen’s Kappa 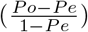 [20], where Po is the observed agreement and Pe is the expected agreement.

#### 2.4.2 System evaluation

We evaluated the three versions of Vitruvius described in Section 2.1.1.3 using the MedQA dataset. Dueto computational constraints associated with testing all versions on the full set of 1,273 questions, we implemented a two-phase evaluation system. In Phase One, all three versions were assessed using a random subsample of 288 questions (23% of the dataset). We progressively scaled and monitored that yielded the best results during this phase. Based on their performance, Versions 2 and 3 advanced to Phase Two, where they were evaluated on the entire dataset. The MedQA dataset offers multiple-choice answers; A, B, C or D. This was the standard of reference used for our model’s evaluation. Correct or standard answers are also known in Machine Learning (ML) as ground truths. Therefore, we adjusted the orchestrator (LLM 1) to produce an answer using the same nomenclature (A, B, C, or D) for direct comparison.

## 3 Results

### 3.1 Dataset distribution

The MedQA dataset includes two types of questions: type 1, which requires one step of reasoning, and type 2, which requires multiple steps. To understand the scope of the dataset, we classified the 1273 questions from the test set according to their medical speciality; the results are displayed in Figure 6. Paediatrics, endocrinology, and haematology/oncology were the specialities with the most questions, with 124 (9.7%), 107 (9.4%), and 103 (8.1%), respectively. The least represented were internal medicine and molecular biology, with 5 (0.4%) and 3 (0.2%) questions, respectively. In this classification, the category “ internal medicine” refers to questions where no subspecialty was specified. Distribution of the MedQA dataset is graphically represented in Figure 5.

**Figure 5:**
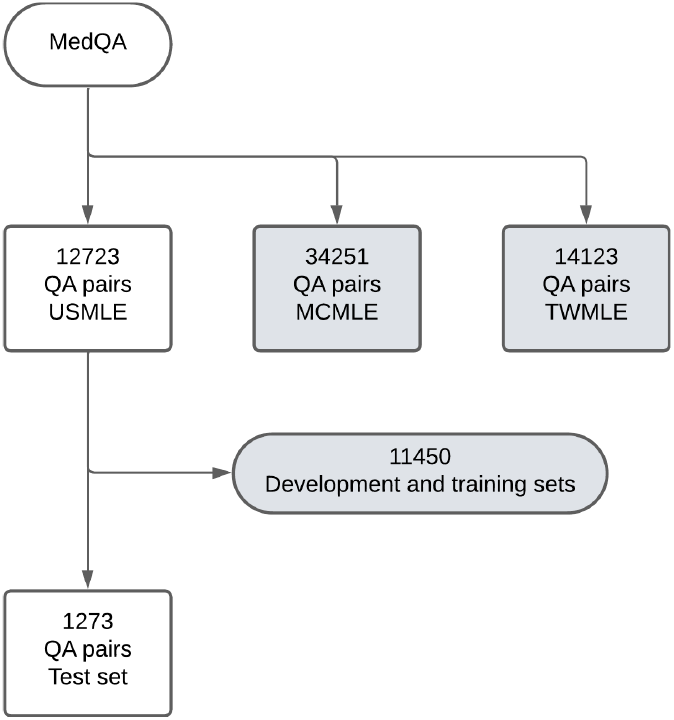
MedQA distribution

**Figure 6:**
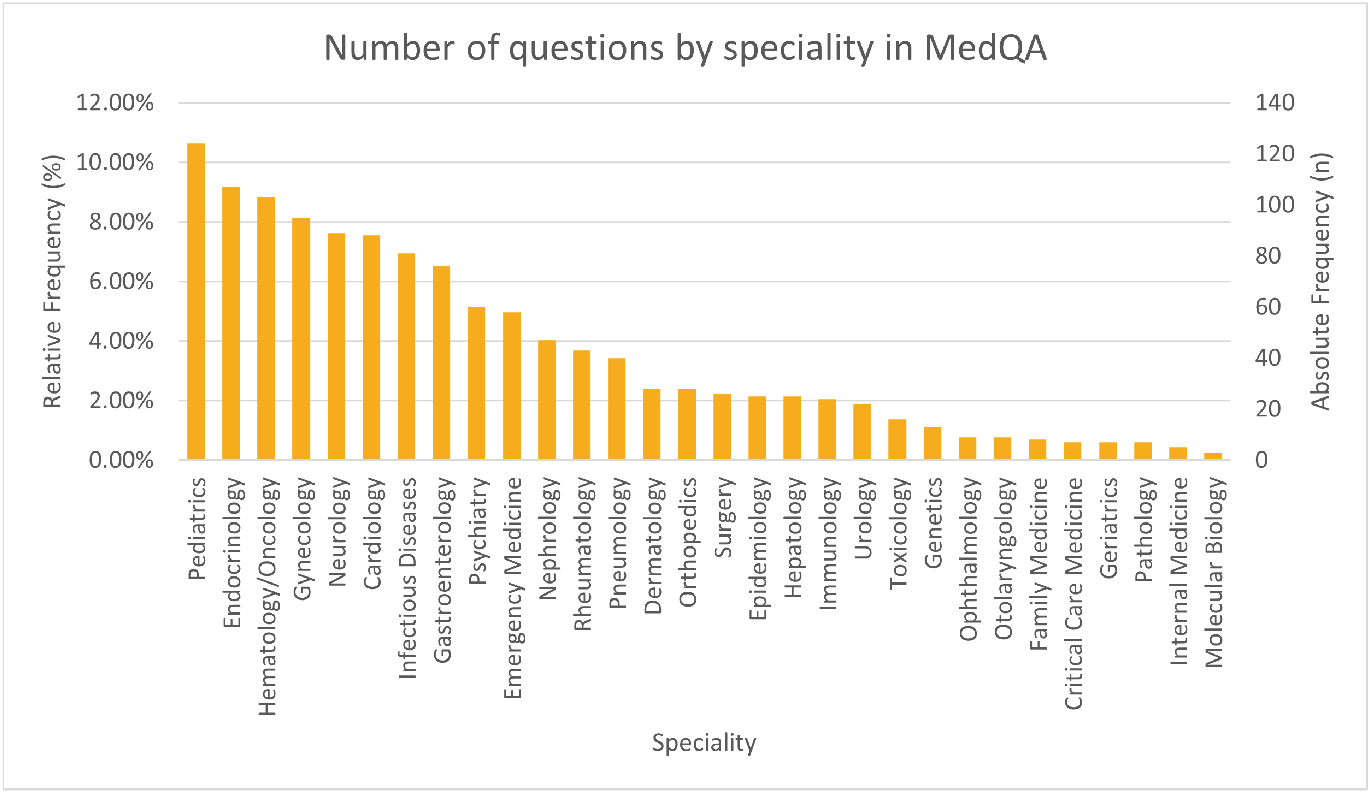
MedQA questions by medical speciality

### 3.2 Development of the model

Following the methodologies outlined in Section 2.1.1.3, we evaluated the three approaches to answer generation using the MedQA dataset. In Phase One, Versions 1 and 2 were tested on a subset of 288 questions from the test set (see Methods section), achieving accuracies of 85.76% and 90.28%, respectively. Based on their superior performance, Versions 2 and 3 advanced to Phase Two, where they were assessed on the complete test set of 1,273 questions. In this phase, they achieved accuracies of 85.15% and 90.26%, respectively. For comparison, accuracy for version 3 on the initial subset (288 questions) was also obtained, achieving 93.06%. These results are summarized in Table 1.

**Table 1:** Accuracy results for the three different versions of Vitruvius.

| Metric | Vitruius |  |  |  |  |
| --- | --- | --- | --- | --- | --- |
|  | Version 1 | Version 2 |  | Version 3 |  |
| Accuracy | 85.76% | 90.28% | 85.15% | 93.06% | 90.26% |
| Correct questions (n) | 247 | 260 | 1084 | 268 | 1149 |
| Total questions (n) | 288 | 288 | 1273 | 288 | 1273 |

#### 3.2.1 Vitruvius version 3

This section presents further results for the final version of Vitruvius that expand on internal and external variability. Table 2 shows the confusion matrix obtained for Vitruvius version 3 compared to the answers provided by MedQA (standard). We assessed internal and external variability using the observed and expected values. The following matrix presents the observed values for the four options in the multiple-choice QAs. Table 3 and Table 4, respectively.

**Table 2:**
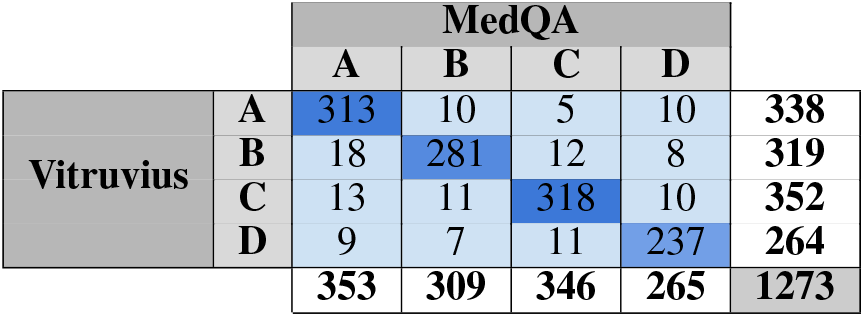
Confusion matrix for the final version of Vitruvius (version 3)

**Table 3:**
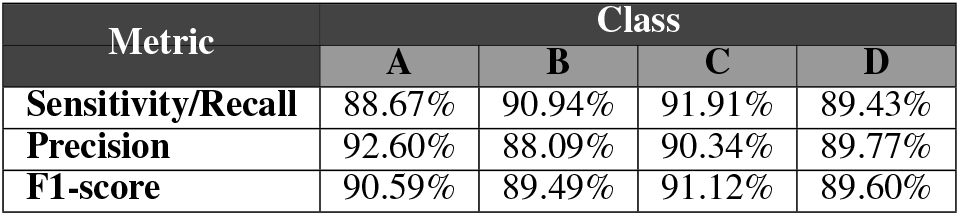
Assessment of sensitivity, precision and F1-score by class (A, B, C and D).

**Table 4:** Comparison to reported State of the art.

| Model | Test set questions used (n) | Accuracy |
| --- | --- | --- |
| **Vitruius v3** | 1273 | **90.26%** |
| **GPT-4o** | 1273 | 87.51% |
| **GPT-4o mini** | 1273 | 73.61% |
| **Med-PaLM 2** | 1273 | 85.40% |
| **GPT-4 (Medprompt)** | 255 | 90.20% |

Sensitivity, precision, and F1 Scores for all four classes showed low variability (Table 3), varying less than 5% across classes for each metric. Sensitivity ranged from 88.67% to 91.91%, precision from 88.09% to 92.60%, and F1-scores from 89.49% to 91.12%. These metrics indicate a balanced model performance across different classes. When comparing the model’s answers to the standard MedQA, we achieved an overall accuracy of 90.26% and a Cohen’s Kappa coefficient of 86.96% (see Table 4), reflecting a high level of agreement.

#### 3.2.2 Model’s classification errors

We manually analyzed a sample of incorrect responses to better identify Vitruvius’s limitations. Figure 7. depicts three of those cases. Figure 7A outlines an ethical case in which a junior surgery resident makes a mistake that is promptly fixed, and thus, the attending says it should not be in the report or disclosed to the patient. According to MedQA the correct answer was B: “Tell the attending that he cannot fail to disclose this mistake”. However, Vitruvius answered C: “Disclose the error to the patient and put it in the operative report”.

**Figure 7:**
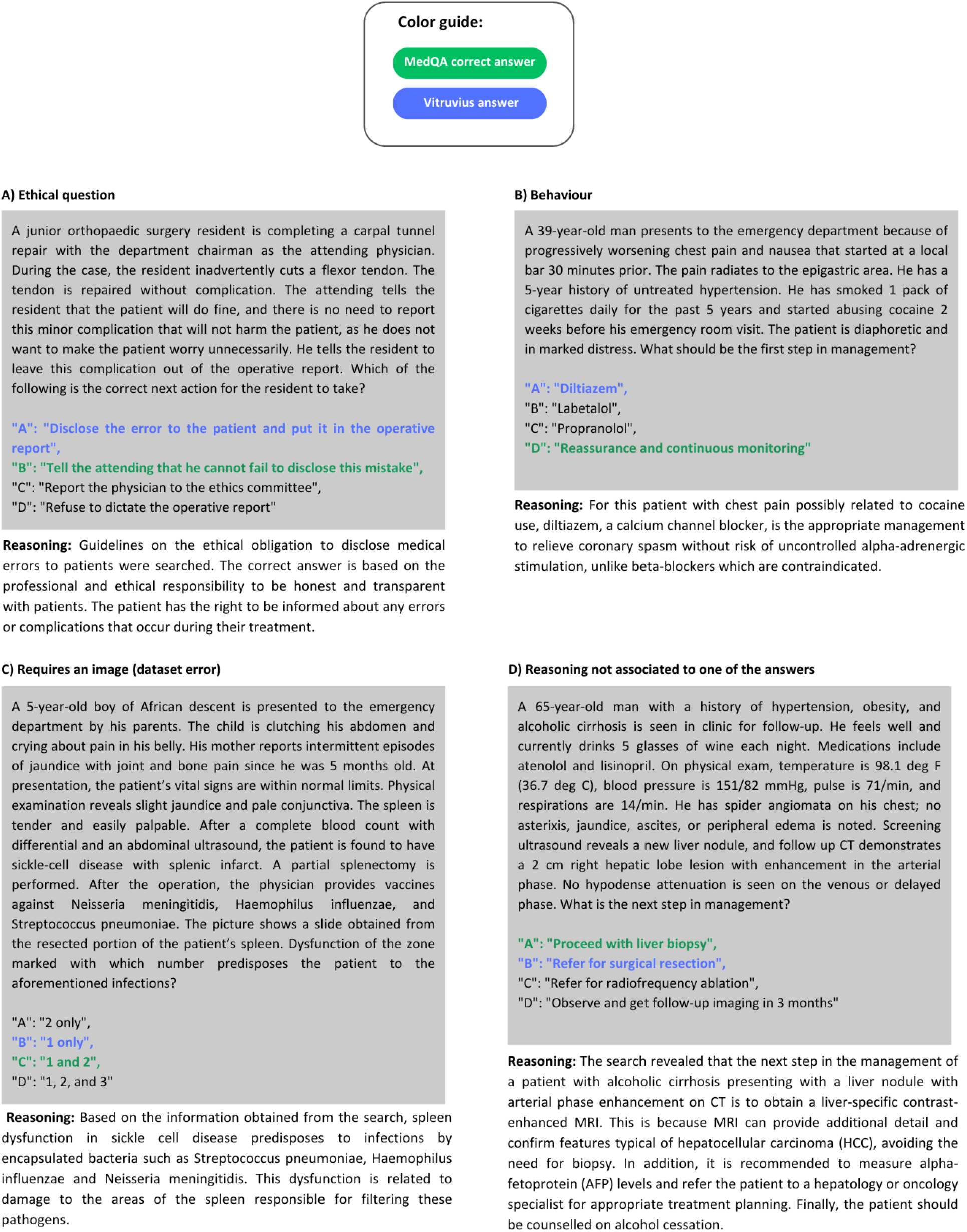
Four examples of incorrect responses. A) a mistake in an ethical question; B) a mistake in a question about behaviour; C) a question that required an image to be answered; D) the Model’s reasoning does not match any of the possible answers

Similarly, Figure 7B shows a question about the first step in managing a patient with chest pain and nausea who is distressed and has consumed cocaine. While our model identifies a possible medication for the patient’s condition (Diltiazem, a calcium-channel blocker used to treat high blood pressure and control chest pain [24]), the correct answer, according to the standard Med-QA, is D: “Reassurance and continuous monitoring”.

Figure 7C shows an apparent error in the benchmark dataset, where the answer to the question requires the analysis of an image (picture of a spleen biopsy). However, no photos or images are present in the dataset. The model attempts to provide an answer by searching for information in the original case and chooses option B: “1 only”. Its reasoning is based on the patient’s clinical history and states the spleen’s main findings in sickle cell disease, the pathology in the clinical case. Finally, Figure 7D shows a case where the model’s reasoning does not match the available options, and the reason for its final answer is unclear. The next section will discuss these cases, especially the latter, as the model’s answer represents a potential risk to the patient’s safety.

### 3.3 Performance compared to SoTA models

Table4 presents the accuracy of Vitruvius Version 3 in comparison to other state-of-the-art models, including GPT-4, GPT-4o mini, Med-PaLM 2 [7], GPT-4 Med prompt [19], and a specialized GPT-4 configuration designed to simulate a hospital environment [20]. The table also includes the number of MedQA questions used to evaluate each model’s performance. Notably, the tests for GPT-4o and GPT-4o mini were conducted by our research team, as a full evaluation of these models on 100% of the MedQA dataset had not been previously published.

## 4 Discussion

This paper introduces the development and initial evaluation of Vitruvius, a system composed of five LLMs designed to answer medical questions in different fields using high-quality clinical research, clinical practice guidelines as sources of information and the search and retrieval capabilities embedded in the LLM architecture.

We evaluated the model’s ability to answer clinical questions using the 1273 QA from the USMLE medical boarding exams in the MedQA dataset. After experimenting with different approaches to obtain the highest possible accuracy compared to other SoTA LLMs, we achieved an accuracy of 90.26% with the final version.

The improvement in accuracy (4.5% compared to Version 2) was achieved by leveraging multiple answers using background knowledge and information retrieved using the search capabilities. Due to LLMs encoding and decoding processes, there is a certain level of randomness in language generation that results in different answers being provided to the same query if asked multiple times. We used this to our advantage by instructing the model to generate three answers from the same context information. This approach resulted in more accurate answers by giving the LLM more chances to include all relevant information and to improve answer consistency. We also added one answer generated from background LLM information (without using RAG) because some reasoning steps required analysis that does not depend on the extracted data. This means that some universal concepts, not necessarily stated or rated in guidelines or research articles, are also needed to answer some questions correctly. For example, questions concerning ethical considerations, those that require the analysis of human behaviour, or those that require mathematical formulas.

Finally, we gave the system the capability to judge and unify all information into one final answer. A paper by Badshah and Sadjjad evaluated LLMs as judges and demonstrated an improvement in accuracy and reliability, establishing a correlation with human judgment [25]. Our experimentation further confirms this, as including an LLM judge led to higher performance evaluation metrics.

Studies evaluating LLMs on MedQA have occasionally reported bias with multiple-choice answers, where the model can produce unbalanced responses by disproportionately selecting or disregarding a specific label [13]. An analysis of internal variability (Tables 2 and 3) shows that the model’s performance is consistent across all categories and metrics, with a maximum variability of 4.52% in precision. This increases our confidence in the model’s retrieval and reasoning abilities.

Vitruvius’ accuracy of 90.26% compares to and surpasses the currently described SoTA of LLMs that used the same number of QAs (1273) as shown in Table 5; for the model Med-PaLM2, the maximum accuracy was 85.4% [11], for GPT-4o accuracy was 87.51%, while GPT-4o mini only showed a 73.61%. When using fewer questions, GPT-4 obtained an accuracy of 90.2% on 255 QA pairs (20% of the original database), using a special prompting strategy coined Medprompt [26], and GPT-4 obtained 93.06%, the higher accuracy, but only using 72 QA [27]. Higher marks in human performance are described around 87% of correct answers. The human exam’s actual threshold or passing score equals or exceeds 60%. Additionally, one study reported an expert score of 87% for the USMLE [13].

The results highlight Vitruvius’ excellent performance, demonstrating its ability to retrieve relevant information and apply reasoning skills. These capabilities position it above the current state-of-the-art LLMs evaluated on this dataset, including our validation of GPT-4o, which achieved an accuracy of 87.51%.

Nevertheless, a physician’s rigorous evaluation of the suggested answers of the model is not optional. Figure 7D showed a case of an incorrect response, which demonstrated faulty reasoning and an answer that could directly harm the patient. Here, Vitruvius selected option B: “Refer for surgical resection”, while the standard database recommended option A: “Proceed with liver biopsy”. This example represents one of the more severe consequences of incorrect responses from IA models. Hendrycks et al., from the Centre for AI Safety, published in October 2023, “An Overview of Catastrophic AI Risks”, a review and classification of the different risks that AI technologies could pose, state that rigorous safety culture can reduce the probability of accidents [28]. In the medical field, safety practices include the close supervision of AI tools. Still, were-state that the purpose of Vitruvius is to provide health personnel with a personal medical knowledge and research assistant, which can be used as source of information and quality search engine rather than to make unsupervised decisions in patient care.

Another aspect to evaluate in the example of Figure 7D is that, in this case, the search provides information for reasoning that does not match any of the given answers. The model’s reasoning indicates that the next step is a liver-specific contrast-enhanced MRI. However, this is not among the options, and it is unclear why the model chooses option B. For Vitruvius, a closed set of possible answers limits the ability to assess the model’s reasoning beyond the given options. This model is designed to answer open questions, which is why there is a possibility that its reasoning does not match the answer options. It would be interesting to analyze it using a different prompting strategy that explicitly instructs the model to examine the four options before retrieving the relevant information. This way, it would be instructed to make an improved search query and find the appropriate information to provide an answer within the four options’ limits. Additionally, in a real interaction the user would be able to make follow up questions or create instructions that could guide the model in its analysis.

Clinical cases that need universally accepted truths to be adequately assessed are another issue in our critical assessment of the model. These include human behaviour, ethics, and appropriate behaviour, which clinical guidelines or research articles do not usually explain. This is the case for the clinical question in Figure 7A, where the model must analyze the best first step to manage an ethical dilemma. In this case, all possible answers demonstrate ethically correct actions where the patient’s right to be informed is respected, as is evidenced in its reasoning. Nonetheless, the correct answer lies in a more subtle analysis of the first course of action by considering the hierarchy between an attending physician and a resident (student). These subtleties of human interaction and preferences are sometimes complex for the model to understand and represent limitations.

The example in Figure 7B shows another side of this same limitation. In this case, the question is, “ What should be the first step in management?”. Vitruvius’s medication analysis is partially correct since the use of Beta-blockers such as labetalol (B) and propranolol (C) are contraindicated in cocaine-associated chest pain. On the other hand, Diltiazem (A), Vtrivius’s answer, is not contraindicated but is not recommended as the first line of treatment [29]. The correct answer is D, “ Reassure and continuously monitor”, which considers an approach that needs some common sense without signs or symptoms pointing to immediate pharmacological intervention.

On the other hand, the example in Figure 7C shows a limitation of the reference dataset as it fails to provide the model with the complete information it needs to respond. This highlights the importance of further validating the model with different datasets and shows that these can contain mistakes.

It is necessary to involve clinicians in evaluating these tools to provide a correct assessment of its benefits, limitations and areas of improvement. Real-life interaction of human and machine should give further insight into the model’s strengths and weaknesses and guide algorithm improvements. In our next manuscript, we aim to further validate Vitruvius by testing it on a wider range of datasets, including questions from additional sources, and conducting evaluations with clinicians to gather direct user feedback. We will also assess its performance across various subtasks to evaluate its generalizability. Additionally, we will compare Vitruvius’ impact on productivity and usefulness against current medical search engines.

## 5 Conclusions

Vitruvius demonstrates robust clinical comprehension and reasoning capabilities, effectively retrieving and answering questions across a wide range of medical topics. Compared to other models tested on the MedQA dataset, Vitruvius achieves state-of-the-art performance, exceeding the current reported best accuracy by 5% (Med-PaLM 2 [7]). Vitruvius also surpasses the foundational models we tested, increasing accuracy by 3% compared to GPT-4o. Future investigations will test Vitruvius in different QA datasets that pose other challenges and directly with clinicians for usefulness in real clinical practice. Nevertheless, Vitruvius is designed as an educational research tool without and does not intend to replace human judgment. While it enhances productivity and aids healthcare professionals, it should not be utilized by individuals unqualified to reason with medical data. Vitruvius aims to augment the expertise of clinicians, ensuring that critical decision-making remains in the hands of trained professionals.

## Data Availability

All data produced in the present study are available upon reasonable request to the authors

## 6 Contributions and competing interests

MCV Machine learning engineer:

Methodology, software, validation

IL Biomedical engineer:

Research, validation, manuscript writing

NCV Medical epidemiologist:

Research, validation, manuscript writing and editing

JM Machine learning engineer:

Methodology, project administration

MFG Sales Development Representative:

Methodology, project administration, research

JZ CEO and Co-founder:

Supervision

LV President and Co-founder:

Supervision

